# The validity and reliability of parent’s recall for routine Immunization in Cameroon: an evaluative study

**DOI:** 10.1101/2022.02.17.22271070

**Authors:** Martin Ndinakie Yakum, Atanga Desmond Funwie, Atem Bethel Ajong, Zahir Shah

## Abstract

**Introduction:** In the absence of immunization documentations, parent’s recall is used to assess children immunization status. During the 2018 demograhpic and health survey in Cameroon, parent’s recall was the only source of information for 47% of chidren assessed for immunization coverage. The objective of this study was to determine the validity of parent’s recall for immunization using the vaccination card as the reference in Yaounde-Cameroon.

**Methods:** The study targeting parents of children aged 0-59months who had their children’s vaccination cards. The immunization history of each child was taken based on both parent’s recall and vaccination card. Using the vaccination card as a reference, the sensitivity, specificity, positive predictive value and negative predictive value of parent’s recall were calculated. The degree of agreement and the kappa statistics between the two methods were calculated using R version 4.1.0 (2021-05-18).

**Results:** A total of 529 households were visited and 87 elligible parents enrolled. Approximately 55.2% of the children were girls and 53% of them were aged 12-59 months. In total, 94.25% of the participants enrolled were one of the biological parents of the children, with mothers making the majority 86.20% of participants. When combined for all vaccines in the EPI (i.e. one dose BCG, 4 doses of OPV, 3 doses of pentavalent vaccine, 3 doses of PCV-13, 2 doses of rotavirus vaccine, one dose of measles/rubella vaccine and one dose of the yellow fever vaccine), the sensitivity, specificity, positive predictive value, and negative predictive value of parent’s recall were 63%, 60%, 90%, and 23% respectively. The degree of agreement between the two sources was highest for BCG(94%) and lowest with Polio2(32%). Parent’s recall(94%) was most likely to correctly predict BCG vaccination status of a child than using the scars on the forarm(74%).

**Conclusion:** Our conclusion is that validity and reliability of parent’s recall vary a lot across different vaccines and parent’s recall is not very reliable for immunization status assessment in children. Parent’s recall is preferred for verifying BCG immunization to scars on the forarm. In general, we recommend that parent’s recall for routine immunization should be used only as a last resort or for BCG, and measles and Yellow Fever vaccines.

## 1. INTRODUCTION

Parent’s recall for immunization can be defined as the ascertaining of children immunization history based solely on the parent’s/guardian’s declaration without any documented proof [1]. During immunization service delivery, the health provider checks the immunization history of the child and identifies vaccines that are due or missed with respect to the child’s age. In the absence of any document to prove the real vaccination status of the child, the provider interviews the child’s parents or guardians in order to determine the child’s immunization status[2]. In the same way, researchers equally rely on parent’s recall when the vaccination card is not available to evaluate the vaccination status of a child enrolled in survey [3].

The use of parent’s recall as a source of information on children immunization various across countries depending on organization and accessibility of immunization information system[4]. In Cameroon, investigators relied on parent’s recall during immunization surveys for 30%-70% of children enrolled[5–8]. The case was different in other context, 3% in Tripura[9], 67% in Pakistan[8], 5% in Tanzania[10]. In Cameroon, immunization data are registered in paper-based registers, stored at the level of the health faciolity and individual vaccination cards, stored at the level of household[11]. However, the maintenance of the vaccination register is generally very poor and sometimes not updated[12,13].

Though parent’s recall is used the last resort to assess the immunization status of children, it is kwon that data collected through parent’s recall does not always match with the real immunization history of the child[14,15]. In the first place, the parent/guardian accompanying the child might not be the same person who was taking care of the child in the past. This can be the case if the biological parents of the child died at some point or unable to accompany the child because of occupations or illness[16]. Secondly, the parent’s recall might be incorrect simple because the parent partially or fully forgot the immunization history of the child in question[17]. Lastly, because the investigator relies on parent’s recall, the parent could intentionally decide to give incorrect information and there will be no way to verify[18].

A few number of studies have assessed the validity of parent’s recall for immunization using vaccination card or vaccination register as the gold standard in some countries[4,16,19,20]. Based on the findings from these studies, the specificity, sensitivity of parent ‘s recall for immunization varies across contexts and vaccines[15,16].

A systematic review on the validity of parent’s recall observed that studies in the subject matter were very few in low-middle income countries(11%) where investigators rely very largely on household information for immunization history assessment[16]. The study concluded that there is no enough evidence to make a definitive conclusion on the subject[16]. No study has been done in Cameroon to assess the context specific situation. The objective of this study was to determine the validity of parent’s recall for routine immunization in Cameroon using vaccination card as the reference.

## 2. MATERIALS AND METHODS

### 2.1. Ethical Approval

This study was authorized by the regional ethics committee for the center region of Cameroon with the authorization reference: No: 01410/CRERSHC/2021. All potential paticipants were well informed about the study objective and procedures of data collection by the study investigators. Before consenting, potential participants were given the chance to ask questions for clarifications and they were free to accept or refuse participantion without any influence or consequence. Verbal informed consent was obtained from all participants before enrollment. The consent for children participants was obtained verbally from their parents after they were properly informed by the investigators. As explained to the participant during information process, participants who consented to participate were equally free to withraw at anytime without having to explain their decision.

### 2.2. Research design

This was an evaluative study targeting parents of children aged 0-59months who had their children’s routine vaccination cards. The immunization status of each child was recorded based on parent’s recall and compared with the information from the vaccination card(reference sources) to estimate sensitivity, specificity, positive predictive value and negative predictive value of parent’s recall. Data were collected through a household survey in which participants were interviewed and vaccination cards verified. However, with the acknowledgement that parent’s recall is useful rather to the population subgroup without a vaccination card, we compared the parent’s recall ability for BCG vaccine between children with cards and those without card, using BCG scars on the forarm as reference, for quality checks. The reliability of parent’s recall was estimated using Kappa statistics and degree of agreement between the two sources of information. Data were analysed with R version 4.1.0 (2021-05-18).

### 2.3. Research area

This study was done in six(6) health districts in Cameroon: Biyem assi, Cite verte, Djoungolo, Efoulan, Nkolbisson, and Nkolndongo. The study area was Yaoundé Cameroon.

### 2.4. Study population

This study targeted parents (or guardians) of children under five years, living in Yaounde that were in possession of their vaccination cards. All potential participants who could not present the vaccination card of their children were excluded from the analysis. However, aprticipants without card were used for quality check analysis.

### 2.5. Sample size calculation

Sample size needed for this study was calculated using the formula for sensitivity study[21]. The parameters used for the sample size estimation included the following: expected sensitivity of 93.4%[10], Zα/2 at 95% confidence interval 1.96, expected vaccination coverage of 42%[3], and the desired precision of 9%. We obtained a sample size of 101 participants. When we considered the vaccination card retention in Cameroon (57%), average household size(4.9), and proportion of children under five years in the population[3], we estimated to interview 529 households in order to obtain the desired sample size.

### 2.6. Sampling Methods

Household selection in the field was done using a 2-stage cluster sampling. A total of 30 clusters constituting of 24 households each were assessed. Clusters were selected with probability proportionate to population size (PPS) and households within cluster selected by restricted sampling. The restricted sampling here refers to a modified form of systematic sampling in which instead of using sampling interval in a systematic way, we randomly selected one household within successive sampling interval. The sampling interval in check cluster was slightly different depending on cluster size. A total of 24 households were selected and assessed in each cluster. This method was preferred to give more room for chance factor in household selection.

### 2.7. Data collection

The data collection tool used in this survey was the questionnaire used by demographic health survey in Cameroon in 2018 for immunization coverage[3]. However, unlike DHS in which parent’s recall was used in the absence of vaccination card, we used both sources at the same time for all participants. Data collection tool was designed in KoBo toolbox and deployed in tablets for electronic data collection. Prior to data collection, data collectors were trained and tools pretested.

### 2.8. Data management and data analysis

Data analysis was done with R version 4.1.0 (2021-05-18). Using vaccination card as our reference source, we calculated sensitivity(se), specificity(se), positive predictive values (PPV) and negative predictive values (NPV) of parent’s recall with their corresponding 95% confidence interval (CI). These values were calculated per vaccine dose and for the all EPI vaccines combined(i.e. one dose BCG, 4 doses of OPV, 3 doses of pentavalent vaccine, 3 doses of PCV-13, 2 doses of rotavirus vaccine, one dose of measles/rubella vaccine and one dose of the yellow fever vaccine). Besides, we calculated the degree of agreement between the 2 methods and the reliability of the test estimated using Kappa statistics. These values were also calculated per vaccine dose and for the all vaccine combined.

As a control check, we compared parent’s recall ability for BCG vaccine between children with vaccination cards and children without cards. This was to evaluate if the recall ability between the two groups significantly differ, and hence would help in the interpretation of our findings.

## 3. Results

### 3.1. Sample description

A total of 529 households were assessed and 304 children aged 0-59 months identified of which 87(24%) had their vaccination cards and 217(76%) without vaccination cards. Table 1 presents the age and sex distribution of the children with card whose parents were enrolled and children without card, not enrolled into the study. Approximately 55.2% of the children with cards were girls and 47% of them were aged 0-11 months. In total, 82(94.25%) of the participants enrolled were one of the biological parents of the children with mothers making the majority 75(86.20%) of participants.

**Table 1:**
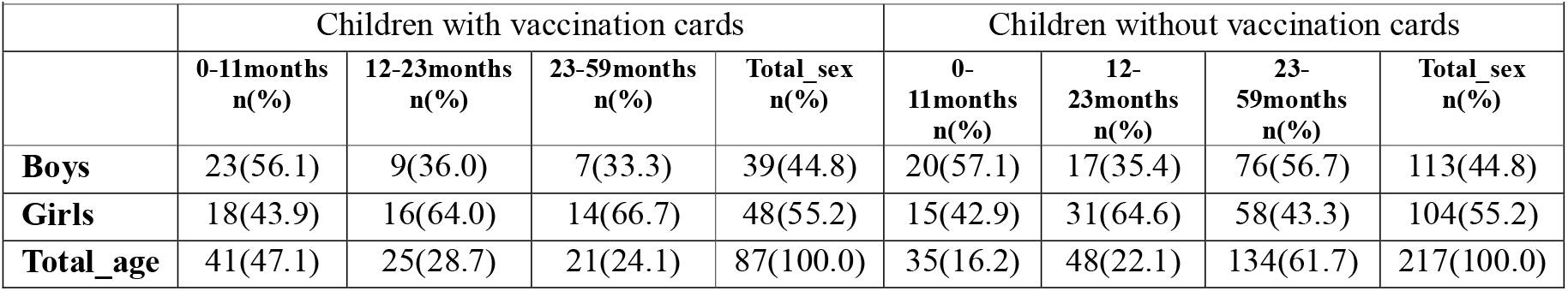
age and sex distribution of children whose parents were enrolled for parent’s recall study in Yaounde

|  |  |  |  |  |  |  |  |  |
| --- | --- | --- | --- | --- | --- | --- | --- | --- |
|  | Children with vaccination cards |  |  |  | Children without vaccination cards |  |  |  |
|  | 0-11months<br>n(%) | 12-23months<br>n(%) | 23-59months<br>n(%) | Total_sex<br>n(%) | 0-11months<br>n(%) | 12-23months<br>n(%) | 23-59months<br>n(%) | Total_sex<br>n(%) |
| **Boys** | 23(56.1) | 9(36.0) | 7(33.3) | 39(44.8) | 20(57.1) | 17(35.4) | 76(56.7) | 113(44.8) |
| **Girls** | 18(43.9) | 16(64.0) | 14(66.7) | 48(55.2) | 15(42.9) | 31(64.6) | 58(43.3) | 104(55.2) |
| **Total_age** | 41(47.1) | 25(28.7) | 21(24.1) | 87(100.0) | 35(16.2) | 48(22.1) | 134(61.7) | 217(100.0) |

### 3.2. parent’s recall ability between children with cards and children without cards

Table 2 compares parent’s recall for BCG vaccine in children with cards and children without cards, using BCG scars on the forarm as the refernce. The results show that parent’s recall ability does not significantly differ bween children having their vaccination cards and children without vaccination card. This finding, suggest that though, our study is conducted in children with vaccination cards, our findings and conclusion can be applied to children without vaccination cards.

**Table 2:** comparison of parent’s recall ability for BCG vaccine between children with cards and children without cards, using BCG scars as the reference

|  | Children with cards | Children without cards | p-value |
| --- | --- | --- | --- |
| Sensitivity | 0.77(0.66, 0.85) | 0.92(0.87,0.95) | 0.8993 |
| Specificity | 0.40(0.05, 0.85) | 0.31(0.11, 0.59) | 0.9026 |
| positive predictive value | 0.95(0.87, 0.99) | 0.94(0.90, 0.97) | 0.9935 |
| negative predictive value | 0.10(0.01, 0.30) | 0.24(0.08, 0.47) | 0.8052 |
| correct classification proportion | 0.75(0.64, 0.83) | 0.88(0.82, 0.92) | 0.9109 |

### 3.3. validity of parent’s recall

When combined for all EPI vaccine doses assessed, the sensitivity and specificity of parent’s recall were 63% and 60% respectively. Also, the positive predictive value and negative predictive value were 90% and 23% respectively. However, the kappa test of agreement shows that parent’s recall is not very reliable. Table 2 shows the number of times parent’s recall was either in agreement or disagreement with the information from the vaccination cards. Note that though only 87 participants were enrolled, depending on the age of the child, one parent could answer up to 15 times on one child, corresponding to the different vaccine doses. This gives rise to the data in table 3 and hence table 4 which presents the validity and reliability parameters of parent’s recall for all vaccines.

**Table 3:**
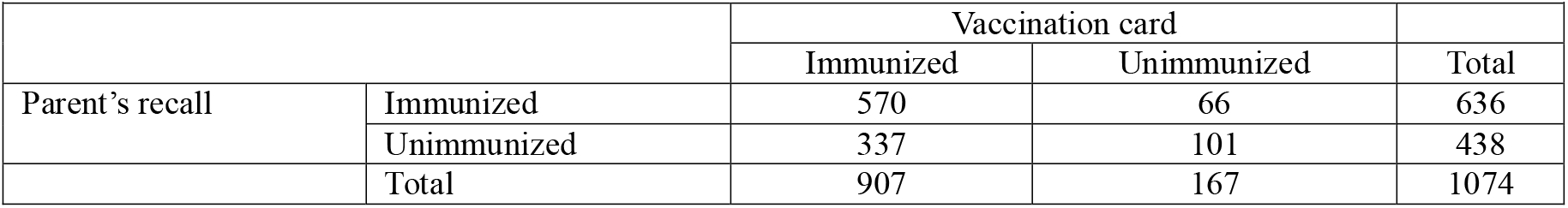
data on immunization history of children obtained from parent’s recall and vaccination cards

**Table 4:** validity and reliability of parent’s recall for all EPI vaccines

| Parameter | Value | 95%CI |
| --- | --- | --- |
| Sensitivity (Se) | 0.63 | [0.60, 0.66] |
| Specificity(Sp) | 0.60 | [0.53, 0.68] |
| Positive Predictive value(PPV) | 0.90 | [0.87, 0.92] |
| Negative Predictive Value(NPV) | 0.23 | [0.19, 0.27] |
| Degree of agreement (d.a) | 0.62 | [0.60, 0.65] |
| Kappa statistics | 0.14 | [0.09, 0.19] |

Table 4 presents the sensitivity, specificity, positive predictive value and negative predictive value of parents recall with their corresponding 95% CIs calculated from the data in table 2. All EPI vaccines refers to one dose BCG, 4 doses of OPV, 3 doses of pentavalent vaccine, 3 doses of PCV-13, 2 doses of rotavirus vaccine, one dose of measles/rubella vaccine and one dose of the yellow fever vaccine

Table 5 shows the parameters of parent’s recall validity and reliability for different vaccines. The validity and reliability parameters of parent’s recall vary a lot across different vaccine doses. Our findings suggest that parent’s recall is more sensitive and less specific for vaccines administered at birth(BCG and OPV0) and vaccines administered at 9 months (Measles and Yellow Fever). When checking the scars on the forearm for BCG compared to vaccination card, the results showed that parent’s recall(d.a=94%) is more reliable than scars(d.a=74%) were very similar to that’s of the parent’s recall for BCG(see table 4). On the other hand, for vaccines administered within 6 weeks-14 weeks, parent’s recall turns to be more specific and less sensitive as shown on table 5.

**Table 5:** validity and reliability of parent’s recall for routine immunization of children per vaccine dose and BCG scars at the forearm using vaccination card as the gold standard.

| **Vaccine** | **Sensitivity(PPV)** | **Specificity(NPV)** | **PPV(Sp)** | **NPV(Se)** | **d.a** | **Ka** |
| --- | --- | --- | --- | --- | --- | --- |
| **BCG** | 0.98 (0.91, 1.00) | 0.50 (0.12, 0.88) | 0.96 (0.90, 0.99) | 0.60 (0.15, 0.95) | 0.94 | 0.51 |
| **Polio0** | 0.91 (0.83, 0.96) | 0.43 (0.10, 0.82) | 0.95 (0.87, 0.99) | 0.30 (0.07, 0.65) | 0.87 | 0.28 |
| **Polio1** | 1.00 (0.84, 1.00) | 0.11 (0.04, 0.21) | 0.26 (0.17, 0.37) | 1.00 (0.59, 1.00) | 0.32 | 0.05 |
| **Penta1** | 0.93 (0.84, 0.98) | 0.62 (0.24, 0.91) | 0.96 (0.87, 0.99) | 0.50 (0.19, 0.81) | 0.90 | 0.50 |
| **PCV-13 1** | 0.46 (0.19, 0.75) | 0.89 (0.79, 0.95) | 0.46 (0.19, 0.75) | 0.89 (0.79, 0.95) | 0.82 | 0.35 |
| **Rota1** | 0.43 (0.18, 0.71) | 0.83 (0.71, 0.91) | 0.35 (0.14, 0.62) | 0.87 (0.75, 0.94) | 0.75 | 0.23 |
| **Polio2** | 0.25 (0.15, 0.38) | 1.00 (0.54, 1.00) | 1.00 (0.79, 1.00) | 0.11 (0.04, 0.23) | 0.32 | 0.06 |
| **Penta2** | 0.43 (0.30, 0.56) | 1.00 (0.54, 1.00) | 1.00 (0.87, 1.00) | 0.14 (0.05, 0.29) | 0.48 | 0.11 |
| **PCV-13 2** | 0.32 (0.20, 0.45) | 0.83 (0.52, 0.98) | 0.90 (0.68, 0.99) | 0.20 (0.10, 0.34) | 0.41 | 0.07 |
| **Rota2** | 0.64 (0.50, 0.76) | 0.43 (0.18, 0.71) | 0.81 (0.67, 0.92) | 0.23 (0.09, 0.44) | 0.59 | 0.05 |
| **Polio3** | 0.28 (0.16, 0.42) | 0.92 (0.62, 1.00) | 0.94 (0.70, 1.00) | 0.22 (0.12, 0.36) | 0.39 | 0.09 |
| **Penta3** | 0.42 (0.29, 0.56) | 0.73 (0.39, 0.94) | 0.88 (0.70, 0.98) | 0.20 (0.09, 0.36) | 0.47 | 0.07 |
| **PCV-13 3** | 0.40 (0.27, 0.55) | 0.64 (0.35, 0.87) | 0.81 (0.61, 0.93) | 0.22 (0.11, 0.38) | 0.45 | 0.03 |
| **MR** | 0.92 (0.78, 0.98) | 0.41 (0.18, 0.67) | 0.77 (0.62, 0.89) | 0.70 (0.35, 0.93) | 0.76 | 0.37 |
| **YF** | 0.95 (0.82, 0.99) | 0.47 (0.23, 0.72) | 0.80 (0.65, | 0.80 (0.44, | 0.80 | 0.47 |
|  |  |  | 0.90) | 0.97) |  |  |
| **BCG Scars** | 0.77 (0.66, 0.85) | 0.33 (0.04, 0.78) | 0.94 (0.85, | 0.10 (0.01, | 0.74 | 0.05 |
|  |  |  | 0.98) | 0.30) |  |  |
| *PPV= positive predictive value, NPV= Negative predictive value, d. a = Proportion of agreement between the two methods and ka = Kappa constant for reliability.* |  |  |  |  |  |  |

Parent’s recall is generally having a good PPVs (77% -100%) and less NPVs(11%-80%) for all EPI vaccines except for OPV1, PCV-13 1, and rota1 that presented opposite findings. However, for MR and YF vaccines, the PPVs and NPVs were similar. In general, parent’s recall was not very reliable with the kappa statistics ≤5% for all vaccines. However, parent’s recall had a good degree of agreement(≥80%) for some vaccine doses such as BCG, OPV0, penta1, pcv-13 1 and YF vaccines.

## 4. Discussions

### 4.1. Summary findings

This study aims to assess the validity and reliability of parent’s recall for routine immunization in children aged 0-59 months in Yaounde-Cameroon. Overall for all vaccines in the EPI (i.e. one dose BCG, 4 doses of OPV, 3 doses of pentavalent vaccine, 3 doses of PCV-13, 2 doses of rotavirus vaccine, one dose of measles/rubella vaccine and one dose of the yellow fever vaccine), the sensitivity, specificity, positive predictive value, and negative predictive value of parent’s recall were 63%, 60%, 90%, and 23% respectively. The degree of agreement between the two sources was highest for BCG(94%) and lowest with Polio2(32%). Parent’s recall(94%) was most likely to correctly predict BCG vaccination status of a child than using the scars on the forarm(74%).

### 4.2. Parent’s recall validity

A few number of studies have assessed the validity of parent’s recall for immunization using vaccination card or vaccination register as the gold standard in a limited number of countries[4,16,19,20]. Based on the findings from this studies, it can be observed that the specificity, sensitivity of parent ‘s recall for immunization various across vaccines[16]. This is similar with our findings as we observed that validity changes with vaccine.

A systematic review on the validity of parent’s recall suggested that we do not yet have enough evidence to make a definitive conclusion on the subject[16]. On the other hand, another study in Tanzania suggested that sensitivity of parent’s recall was very good(>93%) and more stable across different vaccines while specificity varies very widely across vaccines between 16%-95%[10]. However, this particular study in Tanzania included only children borne within 12 months to the survey meanwhile our study targeted children 0-59 months. It could be explained by the fact that more than 50% of our participants were children aged 12-59 months giving more chance for the parents to have forgotten the vaccines received. In another study, it was observed that parents mostly report correctly the immunization status of children less than 6 months than older children[15]. We therefore expect our study to have more recall bias compared to this study in Tanzania. Several other studies have reported that parent’s recall is not reliable for evaluating immunization status of children[15,22]. However, studies have not attempted to describe the variability of this across vaccines. Because of recall’s bias, relying on parent’s recall, during routine service delivery exposes the child to the risk of missing some vaccines or being re-vaccinated unnecessarily[15,16].

Currently, parent’s recall sometimes is the last resort and there is no other way to assess the vaccination status of the child especially in low income countries where the health information system is very weak[18,23]. There is therefore the need to improve the immunization information system in Cameroon. This is to reduce how much we rely on parent’s recall which is less reliable.

### 4.3. study limitations

Our study did not include children who could not present their vaccination cards. It should be noted that parent’s recall is used solely in the absence of the vaccination card, because of this, the ideal study would be done rather in children without a card or at least include them. However, this required a reference data source that includes children without cards such as the health facility immunization registers. This was not possible in our context because of poor maintenance of immunization registers in the health facilities, which are often not up-to-date. For this reason, we decided to check the usefulness of our findings by investigating and comparing the parent’s recall ability for BCG vaccine in children with cards and children without cards, using BCG scars on the forarm as the reference. It came outfrom this assessment that the parent’s recall bias among children with cards was not significantly different from recall bias among children without cards. Therefore, parent’s recall validity in this study are closely similar to the validity of parents recall in the entire population, including children without cards.

## CONCLUSIONS

The sensitivity, specificity, positive predictive value, and negative predictive value of parent’s recall for routine immunization in Cameroon are respectively 63%, 60%, 90%, and 23%. Parent’s recall varies from one vaccine to another and it is more sensitive and less specific for vaccines administered at birth(BCG and OPV0) and vaccines administered at 9 months (MR and YF).

When compared to checking the scars on the forearm for BCG, parent’s recall was more reliable in evaluation BCG immunization in children with a recall bias of 6% against 27% for scars. Generally, parent’s recall is not very reliable for assessing a child’s immunization status. Based on these findings, we propose the following recommendations:

- Parent’s recall for routine immunization should be used only in the absence of vaccination card. However, it could be used with less risk of recall bias if we have to assess only the immunization coverage in BCG, Measles, and Yellow Fever vaccines.
- To verify BCG immunization status of the child when the vaccination card is not available, we recommend to use parent’s recall instead of scars on the forarm.
- Further research is needed to assess the other sources of information for routine immunization in Cameroon such as the vaccination register and vaccination card.

## What is already know on this topic

- Immunization data collected from parent’s recall is less reliable compared to data from vaccination cards and vaccination registers.
- Parent’s recall for routine immunization varies from one context to another.

## What this study adds

- Validity of parent’s recall for every single routine immunization vaccine dose
- Compare parent’s recall bias between children with vaccination cards and those without cards

## Data Availability

All relevant data are within the manuscript and its Supporting Information files

## Authors’ contributions

**MNY** conceived the study, designed the study, led data collection and analysis, write the first draft of thr manuscript; **ADF** conceived the study, contributed to study design and manuscript writing; **ABA** contributed to study design and writing of the manuscript; **ZS** conceived the study, contributed to study design and manuscript writing

## Acknowledgements

We are thankful to the data collection team: Miss DOUANLA KOUTIO Ingrid Marcelle, Miss TCHENGO MASSOM THÉRÈSE ZITA, Miss Christelle Bertyl TCHANA MBETBEUM, and Miss Ngueni Letegnou Nancy. Our appreciation equally goes to Professor Djuidje Marceline, providing training venue for data collectors training.

## LIST OF ABBREVIATIONS

Abbreviation: Definition
BCG: Calmette-Guérin Bacillus (vaccine)
CI: Confidence interval
DHS: Demographic Health Survey
DPT-HepB+Hib: Diphtheria, Pertusis, Tetanus and Hepatitis B + Haemophilus Influenzae type b
EPI: Expanded Program of Immunization
Hib: Haemophilus Influenzae type b
Hepb: Hepatitis B vaccine
IPV: Inactivated Polio Vaccine
Ka: Kappa statistics
MR: Measles and Rubella vaccine
NPV: Negative predictive value
OR: Odds ratio
OPV: Oral Polio Vaccine
PCV-13: Pneumococcal Conjugate Vaccine 13
Penta: Diphtheria, Pertusis, Tetanus and Hepatitis B + Haemophilus Influenzae type b
PPV: Positive predictive value
DPT+Hib+HepB: Diphtheria, Pertusis, Tetanus and Hepatitis B + Haemophilus Influenzae type b
d.a: degree agreement
rota: Rotavirus vaccine
Se: Sensitivity
Sp: Specificity
YF: Yellow Fever

## Attachment: STROBE Statement—Checklist of items that should be included in reports of cross-sectional studies

|  | Item No | Recommendation | Page No |
| --- | --- | --- | --- |
| Title and abstract | 1 | (a) Indicate the study's design with a commonly used term in the title or the abstract | 1 |
|  |  | (b) Provide in the abstract an informative and balanced summary of what was done and what was found | 2 |
| Introduction |  |  |  |
| Background/rationale | 2 | Explain the scientific background and rationale for the investigation being reported | 4 |
| Objectives | 3 | State specific objectives, including any prespecified hypotheses | 5 |
| Methods |  |  |  |
| Study design | 4 | Present key elements of study design early in the paper | 6 |
| Setting | 5 | Describe the setting, locations, and relevant dates, including periods of recruitment, exposure, follow-up, and data collection | 6 |
| Participants | 6 | (a) Give the eligibility criteria, and the sources and methods of selection of participants | 7 |
| Variables | 7 | Clearly define all outcomes, exposures, predictors, potential confounders, and effect modifiers. Give diagnostic criteria, if applicable | 8 |
| Data sources/<br>measurement | 8* | For each variable of interest, give sources of data and details of methods of assessment (measurement). Describe comparability of assessment methods if there is more than one group | 7 |
| Bias | 9 | Describe any efforts to address potential sources of bias | 8 |
| Study size | 10 | Explain how the study size was arrived at | 7 |
| Quantitative variables | 11 | Explain how quantitative variables were handled in the analyses. If applicable, describe which groupings were chosen and why | 8 |
| Statistical methods | 12 | (a) Describe all statistical methods, including those used to control for confounding | 8 |
|  |  | (b) Describe any methods used to examine subgroups and interactions | 8 |
|  |  | (c) Explain how missing data were addressed | 8 |
|  |  | (d) If applicable, describe analytical methods taking account of sampling strategy | 7 |
|  |  | (e) Describe any sensitivity analyses | 8 |
| Results |  |  |  |
| Participants | 13* | (a) Report numbers of individuals at each stage of study—eg numbers potentially eligible, examined for eligibility, confirmed eligible, included in the study, completing follow-up, and analysed | 9 |
|  |  | (b) Give reasons for non-participation at each stage | 9 |
|  |  | (c) Consider use of a flow diagram |  |
| Descriptive data | 14* | (a) Give characteristics of study participants (eg demographic, clinical, social) and information on exposures and potential confounders | 9 |
|  |  | (b) Indicate number of participants with missing data for each variable of interest | 9 |
| Outcome data | 15* | Report numbers of outcome events or summary measures | 10 |
| Main results | 16 | (a) Give unadjusted estimates and, if applicable, confounder-adjusted estimates and their precision (eg, 95% confidence interval). Make clear which confounders were adjusted for and why they were included | 10 |
|  |  | (b) Report category boundaries when continuous variables were categorized | 10 |
|  |  | (c) If relevant, consider translating estimates of relative risk into absolute risk for a meaningful time period |  |
| Other analyses | 17 | Report other analyses done—eg analyses of subgroups and interactions, and sensitivity analyses | 10 |
| **Discussion** |  |  |  |
| Key results | 18 | Summarise key results with reference to study objectives | 10 |
| Limitations | 19 | Discuss limitations of the study, taking into account sources of potential bias or imprecision. Discuss both direction and magnitude of any potential bias | 11 |
| Interpretation | 20 | Give a cautious overall interpretation of results considering objectives, limitations, multiplicity of analyses, results from similar studies, and other relevant evidence | 9-10 |
| Generalisability | 21 | Discuss the generalisability (external validity) of the study results | 11 |
| **Other information** |  |  |  |
| Funding | 22 | Give the source of funding and the role of the funders for the present study and, if applicable, for the original study on which the present article is based |  |

